# Wastewater-based epidemiology to enhance public health preparedness and response during large-scale events: experiences from the 2024 Republican and Democratic National Conventions – Milwaukee, WI and Chicago, IL

**DOI:** 10.64898/2026.01.26.26343470

**Authors:** Peter MV DeJonge, Ian Pray, Rachel Poretsky, Martin Shafer, Sandra L McLellan, Alyse Kittner, Colin Korban, Dolores Sanchez Gonzalez, Adam Horton, Modou Lamin Jarju, Chi-Yu Lin, Erin P Newcomer, Hannah J Barbian, Stefan J Green, Bernardo Burbano-Abril, Nathan Kloczko, Megan Rasmussen, Dagmara Antkiewicz, Adélaïde Roguet, Devin Everett, Melissa K Schussman, Veronica McSorley, Peter Ruestow

## Abstract

**Introduction:** Wastewater-based epidemiology (WBE) was implemented at the 2024 Republican and Democratic National Conventions (RNC and DNC, respectively)—two prominent large-scale events, each with estimated attendances of >50,000 persons. In preparation for event monitoring, the Wisconsin and Chicago WBE programs (associated with the RNC and DNC public health response, respectively) developed site-specific monitoring strategies and response plans, prioritized additional pathogens for event surveillance, and further optimized laboratory workflows to ensure rapid daily data reporting to public health. The Chicago program expanded the sewer sampling network to include new locations closer to event venues than previously available. Sampling was also conducted before the events, to establish baselines for endemic pathogens, as well as after each event to monitor for residual community transmission.

**Methods:** Surveillance was expanded from the four respiratory pathogens regularly assessed by both WBE programs (SARS-CoV-2, influenza A, influenza B, respiratory syncytial virus) to include 3 gastrointestinal pathogens (norovirus, *Salmonella enterica*, Shiga toxin-producing *E. coli*). The Wisconsin program also conducted monitoring for the measles, mumps, rubella, and hepatitis A viruses. Wastewater sampling for the RNC was conducted at the community water reclamation facility level, while at the DNC samples were collected from manholes located downstream of the event venues. For both events, WBE data were summarized and contextualized alongside traditional public health surveillance data in daily situation reports.

**Results:** Between the RNC and DNC response, a total of 112 wastewater samples were collected and assayed to provide concentration data on as many as 11 distinct pathogens of interest. Concentration results for the suite of pathogens were available within 12 to 36 hours of sample collection. In each instance when wastewater concentrations exceeded pre-established thresholds for action and flagged as an alert, other sources of contemporaneous public health surveillance information (e.g., clinical data) did not corroborate the WBE findings.

**Conclusion:** Existing WBE infrastructure in two U.S. cities was readily adapted for public health surveillance at two high-profile, large-scale events. Assays for additional event-relevant pathogens were quickly incorporated into routine laboratory workflows and data from wastewater samples were generated and reported with rapid turnaround-time. In considering the unique benefits of wastewater data, WBE results were a valuable supplement to other public health surveillance data in monitoring potential public health threats during these two large-scale events.

## Introduction

Wastewater-based epidemiology (WBE) is a relatively young, though promising, tool used to conduct community and facility level public health surveillance of infectious diseases.^1–3^ The aggregate nature of wastewater data is inherently economical, deidentified, and representative of the community without selection bias from healthcare access or disease severity.^4–7^ WBE has been used to monitor a wide variety of illnesses, including those caused by respiratory and gastrointestinal pathogens.^8–14^ Pathogen trends in wastewater concentration data correlate well with the incidence of corresponding clinical illness observed in the community and WBE data can better reflect disease prevalence when clinical surveillance is sparse.^10,11,15–18^ For some pathogens, wastewater signals may precede that of traditional surveillance data (e.g., hospital admissions, syndromic surveillance) which allows for more timely public health intervention.^16,19,20^

A potentially valuable use case for WBE is during large-scale events, where mass gatherings of attendees—ranging from tens to hundreds of thousands of persons—increase the potential for transmission of infectious diseases.^21–24^ Monitoring disease and illness during mass events via traditional methods is often challenging. For instance, clinical-based surveillance might be compromised when travelers avoid healthcare due to cost, unfamiliar language or customs, or lack of experience with the local healthcare system.^27^ If there is significant asymptomatic spread among a large group of attendees, public health agencies may identify outbreaks only after they have already spread widely. This delay makes it especially difficult to intervene effectively during the critical period for control and especially when large-scale events are of a short duration. For instance, asymptomatic spread during the incubation period of any given disease complicates disease detection in traditional public health surveillance modalities. To address these challenges, the implementation of WBE as a public health tool for pathogen surveillance at large-scale events has been proposed.^28,29^

Previously published results of WBE at large-scale events, most of which were limited to surveillance for one or two pathogens, provide evidence for its value. These efforts included surveillance for SARS-CoV-2 at the 2020 Olympic and Paralympic Games in Japan;^30^ surveillance for mpox virus and SARS-CoV-2 at 2022 Southern Decadence, a Pride Festival in New Orleans, Louisiana;^31^ surveillance for enterovirus, SARS-CoV-2, and poliovirus at the 2022 World Cup in Qatar;^32^ and surveillance for SARS-CoV-2 at the 2022 and 2023 Two Oceans Marathon in South Africa.^33^ One of the broadest applications of wastewater pathogen testing at large-scale events, was conducted during the 2022 World Athletics Championships in Oregon where wastewater was tested for genes associated with SARS-CoV-2, influenza A virus, respiratory syncytial virus, hepatitis A and E viruses, measles virus, and Middle East Respiratory Syndrome coronavirus.^34^ Publicly-available results indicated increased concentrations of SARS-CoV-2 during the Oregon event, with sporadic detections of hepatitis A virus and influenza A virus.

In the summer of 2024, two U.S. cities in neighboring states hosted large-scale events for the two primary U.S. political parties. During July 15–18, 2024, the Republican National Convention (RNC) was held in Milwaukee, Wisconsin. A month later, during August 19–22, 2024, the Democratic National Convention (DNC) was held in Chicago, Illinois. Over 50,000 people were estimated to have participated in each event. Both conventions were designated National Special Security Events by the U.S. federal government given their large size, high-profile nature, and attendance of U.S. government officials and dignitaries. Each convention’s incident management plan integrated WBE as a fundamental element of the public health surveillance strategy. The City of Milwaukee Health Department and Wisconsin Department of Health Services collaboratively managed public health surveillance during the RNC; wastewater collection, laboratory processing, and analysis were performed by academic and public health partners of the Wisconsin Wastewater Monitoring Program (WWMP). Similarly, at the DNC, the Chicago Department of Public Health (CDPH) managed the public health surveillance strategy for the event, with wastewater collection, laboratory processing, and analysis performed by academic and public health partners of the Chicago WBE team (CWBE).

In preparation for the two conventions, both WWMP and CWBE enhanced their existing WBE programs by making several key additions.^15,18,35^ First, both programs incorporated new pathogen targets related to gastrointestinal illnesses given the prevalence of these diseases at large-scale events.^22,23,36,37^ Second, WWMP added four additional pathogens to their WBE program. Third, two new sampling sites in Chicago were added, which were closer to the event venues than the WBE program’s existing sampling sites. This report outlines the integration of WBE into public health surveillance protocols for large-scale events, highlighting the utility and advantages of the resulting data. We also offer recommendations for other jurisdictions considering the adoption of WBE for large-scale events, particularly in light of the numerous high-profile mass-gatherings anticipated across the U.S. in the coming years (e.g., 2026 FIFA World Cup games, 2028 Olympics and Paralympic Games in Los Angeles, CA) where WBE could prove valuable.

## Methods

### Program alignment

The WWMP and CWBE worked closely in the year leading up to the RNC and DNC events to implement broadly similar WBE methods and laboratory assays, which facilitated the exchange of best practices and comparison of findings. Both the WWMP and CWBE decided to monitor wastewater levels of SARS-CoV-2, respiratory syncytial virus (RSV), influenza A and B viruses (IAV and IBV, respectively), norovirus (NoV), *Salmonella enterica*, Shiga toxin-producing *E. coli* (STEC). CWBE additionally monitored for levels of the fecal normalization marker pepper mild mottled virus (PMMoV) and WWMP additionally monitored for measles, mumps, rubella and hepatitis A (HAV) viruses. Both programs began testing collected wastewater samples for all pathogen targets twice-weekly, at least six weeks prior to each event to establish pathogen-specific data baselines. During the week preceding and throughout both conventions, daily sample collection was conducted, with an emphasis on minimizing turnaround time from collection to result. Both the WWMP and CWBE established action plans and communication strategies in the event of elevated or concerning WBE data. During both the DNC and RNC, we assessed trends in daily normalized concentration levels across pathogens relative to baseline levels to determine the need for a public health response; for certain pathogens (described below), detection alone was sufficient to warrant a response.

### Sample collection

Samples were collected prior to, during, and following both conventions: from June 15 through July 31, 2024 for the RNC, and from July 1 through September 6, 2024 for the DNC. For the RNC, samples were collected from 2 community water reclamation facilities (WRF) in Milwaukee, WI (Jones Island and South Shore WRF; Figure 1A). These Milwaukee facilities covered water reclamation for all RNC-related event venues, area hotels, and Milwaukee residents. The primary event venue (Fiserv Forum, 18,000-person capacity) was located in the Jones Island sewershed, along with most RNC-associated hotels. The residential populations served by the Jones Island and South Shore WRFs are approximately 470,000 and 615,000 people, respectively. All wastewater samples collected at the WRFs were 24-hour flow-weighted composites (12:00am to 11:59pm each day) and were delivered to the two WWMP wastewater testing laboratories (Table 1): University of Wisconsin-Milwaukee School of Freshwater Sciences (UWM), Milwaukee, WI (30 minute drive from sample location) and the Wisconsin State Laboratory of Hygiene (WSLH) Madison, WI (2 hour drive from sample location).

**Figure 1:**
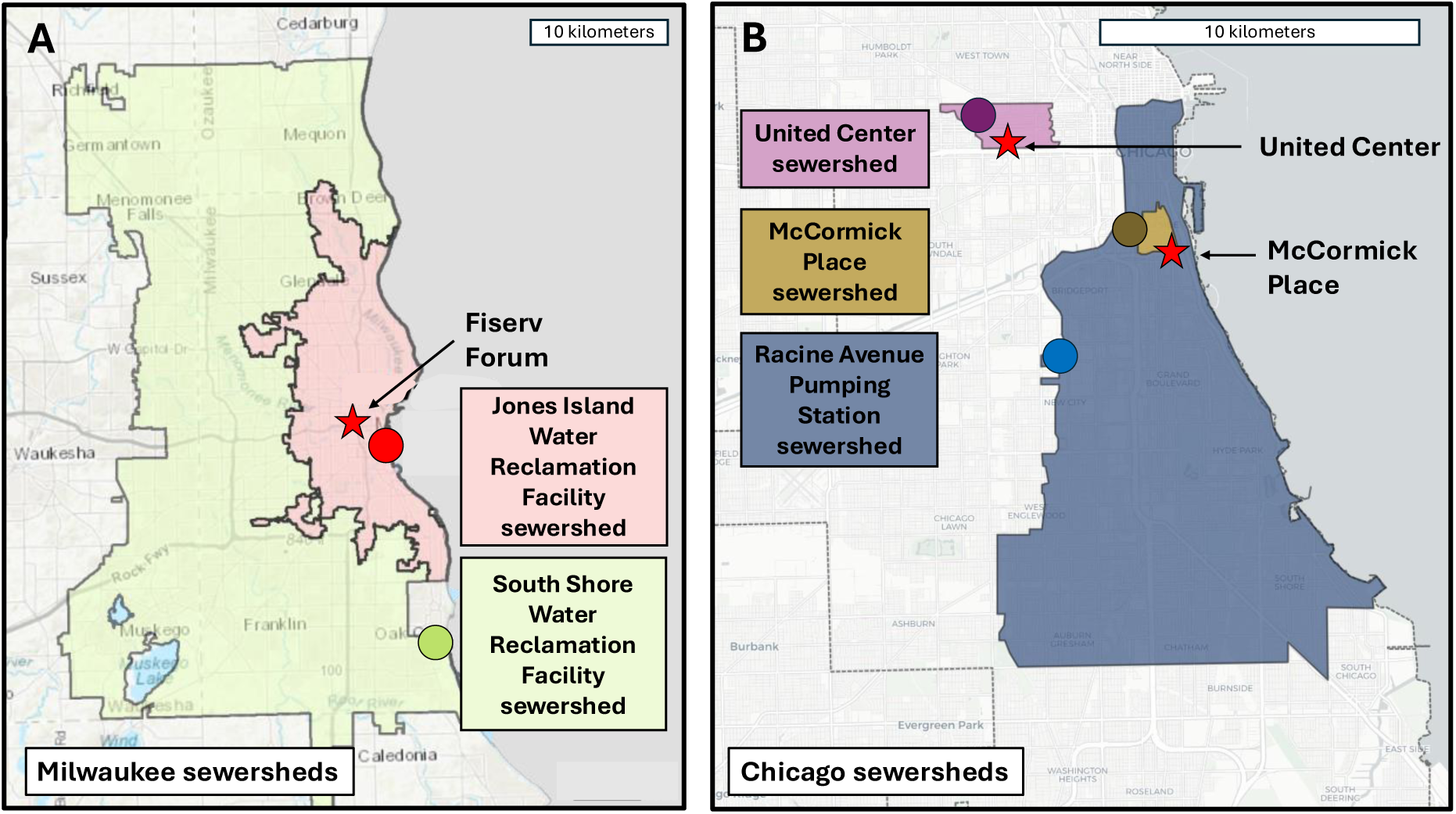
Map of sewersheds monitored by the Wisconsin Wastewater Monitoring Program and the Chicago Wastewater Based Epidemiology team during the 2024 Republican and Democratic National Conventions, respectively. Panel (A) displays the sewershed catchment areas represented by sampling at two water reclamation facilities in Milwaukee, Wisconsin, USA as well as the location of each facility (point) and the main RNC event venue (star). Panel (B) displays the sewershed catchment areas represented by sampling at three manholes in Chicago, Illinois, USA, the location of each manhole (circles) as well as the locations of the two DNC event venues (star).

**Table 1:**
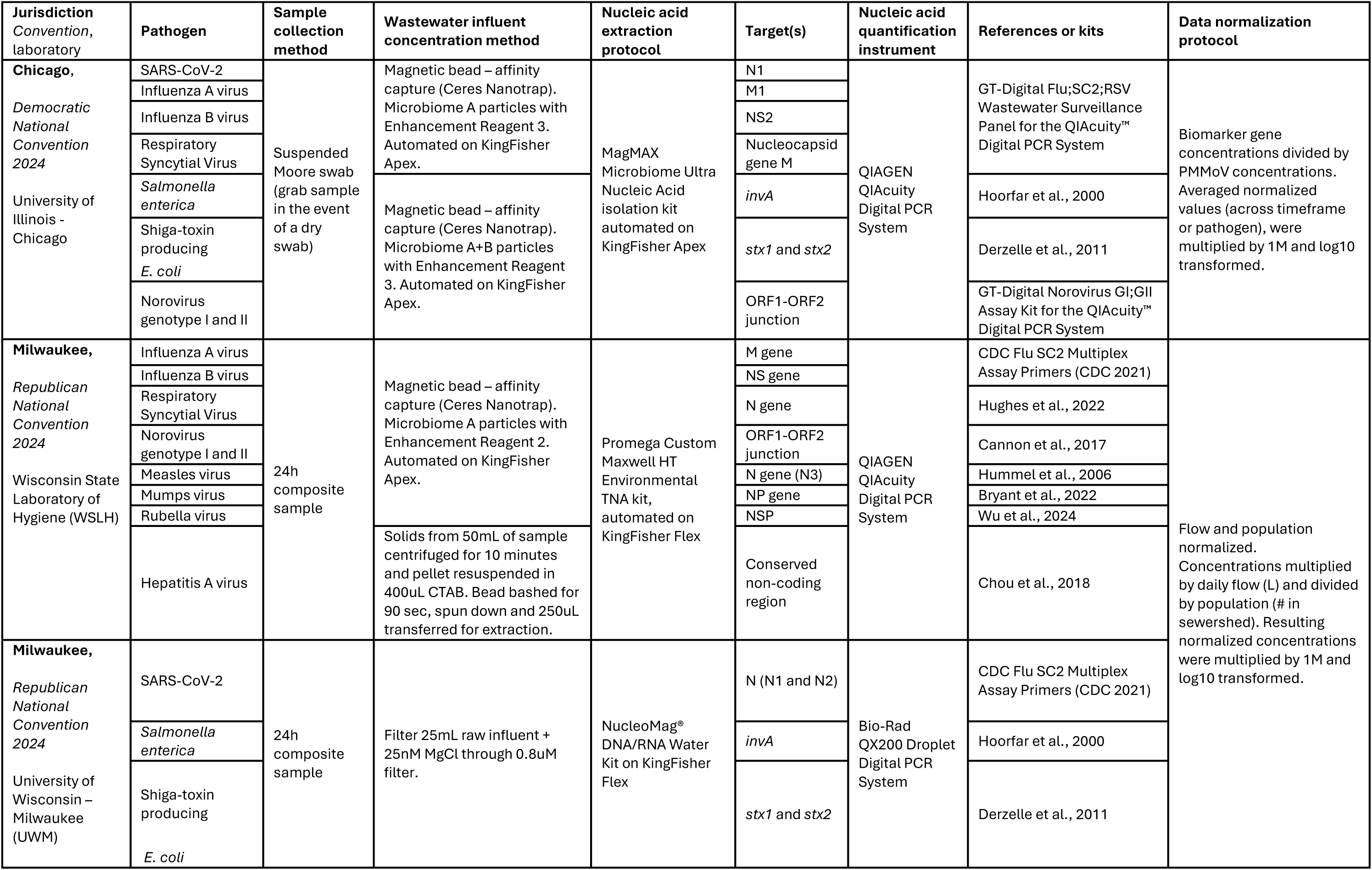
Summary of laboratory and analytic methods implemented at each site.

For the DNC, there were two main venues which hosted event attendees: McCormick Place (18,000-person capacity) and the United Center (21,000-person capacity). Because the single WRF that handles wastewater from both DNC venues serves such a large residential population (approximately 2.5M people), wastewater sampling during the DNC utilized sub-sewershed sampling. Due to security considerations, we could not sample from sewers directly servicing the two venues; instead, two maintenance holes, one outside of each of the venues’ security perimeter, were identified using a hydrodynamic model of the sewersheds where McCormick Center and United Center are located. The model simulated the wastewater flow in the city sewers connected to these buildings and the transport of scalars injected at each one of the venues. The model identified two appropriate locations that captured wastewater from the DNC sites as well as the surrounding communities of 11,210 and 7,870 residents (Figure 1B). Sampling at these two locations began July 1, 2024 and was initially planned through August 23, 2024—the end of the DNC event. During the event, the decision was made to extend sampling at the McCormick Place location through September 6, 2024, to monitor an emergent spike in *S. enterica* concentrations. In addition to the two venue-associated sites, CWBE continued to sample from the other established Chicago sites including the Racine Avenue Pumping Station, a site with a much larger sewershed area (estimated population served: 467,536) that captured the wastewater from many downtown hotels where attendees stayed as well as McCormick Place itself (Figure 1B). All wastewater samples were collected by *Current* (currentwater.org), a CWBE sample collection partner since the fall of 2021. Wastewater was sampled using Moore swabs of sterile cotton gauze suspended in the manholes within stainless steel cages.^38^ These passive samplers were deployed for 24-hours, typically starting at 9:00am each day when the previous sampler was collected. For one sample, the Moore swab yielded an insufficient volume of wastewater due to low water levels that day; as a result, a grab sample—or a discrete collection taken at a single point in time—was utilized that day. Samples were subsequently transported within two hours of collection to the University of Illinois Chicago (UIC) for processing and target quantification.

### Laboratory methods

Pathogens in 10 ml wastewater samples from both the RNC and DNC were concentrated using Ceres® microbiome particles for all targets except HAV, *S. enterica,* STEC, and SARS-CoV-2 in Wisconsin; WWMP samples for HAV testing used solids recovered from centrifugation of 50 ml of wastewater, *S. enterica,* STEC, and SARS-CoV-2 were captured and concentrated using magnesium chloride precipitation followed by HA filtration. WWMP quantified SARS-CoV-2, STEC and *S. enterica* using the Bio-Rad QX200 Droplet Digital PCR System (ddPCR). The other targets were quantified using the QIAcuity™ Digital PCR System (dPCR). CWBE also used dPCR for quantification. Assays used by both WWMP and CWBE are described in Table 1. Additional details regarding the concentration and extraction workflow, PCR protocols, controls, and limits of detection can be found in the Supplementary Material (available upon request).

### WBE data interpretation

For the RNC, wastewater concentrations were normalized by a predefined residential population for each sewershed and the total daily wastewater flow for each sample (Table 1). For the DNC, wastewater concentrations were normalized using the fecal-marker PMMoV, as flow rate was not available in Chicago manholes. For both conventions, all targets were reported individually, except STEC targets (*stx1* and *stx2*) which were combined by calculating the geometric mean of normalized *stx1* and *stx2* concentrations by site by day. For seasonally detected pathogens (IAV, IBV, and RSV) and pathogens not normally present in wastewater (including viruses like measles, mumps, rubella, and HAV), raw wastewater concentrations were compared against the respective limits of detection (LOD) and reported as either detected or not detected. For pathogen targets that were regularly quantified in wastewater (SARS-CoV-2, NoV GI, NoV GII, *S. enterica,* and STEC), daily normalized concentration data were visually assessed for trend and concentration relative to baseline levels using quintiles. The geometric mean of the previous three samples was assigned a category (i.e., *Very Low, Low, Moderate, High, Very High*) based on its value relative to quintile thresholds of baseline data (e.g., 20^th^, 40^th^, 60^th^, 80^th^ percentiles). Notably, in Milwaukee, baseline data was available for >1 year for SARS-CoV-2, RSV, IAV, IBV, NoV GI and GII; however, surveillance for *S. enterica* and STEC only began June 15, 2024, and so WWMP tested archived samples from the 2 WRFs from June through August 2023 for *S. enterica* and STEC. Contrastingly, WBE was entirely new at the two Chicago sites (McCormick and United Center) and only started on July 1, 2024; the amount of baseline data, therefore, was much sparser in for the DNC compared to the RNC.

During each convention, epidemiologists contextualized WBE data alongside more established public health surveillance systems to evaluate the utility of WBE and better inform decision-making. Notable contextual data included the daily volume and type of (i) jurisdictional emergency department visits, (ii) jurisdictional inpatient admissions, (iii) calls to the regional poison center, (iv) jurisdictional ambulance runs, and (v) visits to medical first aid stations onsite at the venues. In Chicago, WBE concentration data from DNC-specific samples was compared to that of samples collected from other WBE sample sites throughout the city. At both the RNC and DNC, epidemiologists discussed WBE results daily and incorporated these data into regular public health surveillance situation reports. Any concerning WBE data signals (i.e., *High* or *Very High* quintile concentrations, persistent increases in trends, detection of rare pathogens) were flagged for additional review.

## Results

### RNC results

During the four-day RNC event, pathogen results were available within 36 hours of sample collection. WBE data elicited several data flags (*High* or *Very High* levels) that were reviewed by the surveillance task force and were included in daily situational reports (Table 2). These data flags occurred for SARS-CoV-2, NoV GII, and STEC for at least one day during the RNC (circled in Figure 2). Quintile flags were compared against other surveillance data (e.g., emergency department visit data) to determine if specific public health actions would be recommended per the response plans. For SARS-CoV-2, the *Moderate* to *High* levels observed were expected and consistent with statewide and national wastewater concentration trends for SARS-CoV-2 leading up to the event. Wastewater flags for SARS-CoV-2 during the event mirrored contemporaneous data from syndromic surveillance—specifically, increases above expected baseline levels in the number of respiratory illness visits and COVID-19 diagnoses in area emergency departments. We noted evidence for a possible increase in community gastrointestinal disease during the RNC; during the event, there were 2 WBE flags based on elevated levels of *S. enterica* and 1 flag for STEC (Figure 2). A day prior to the RNC, we noted 1 wastewater flag for NoV GII. The timing of these WBE flags aligned with increases in syndromic surveillance data for gastrointestinal illness during the event. These concurrent flags from wastewater and syndromic surveillance were reported to stakeholders on the situational report, no specific public health actions were recommended. All other pathogens remained either undetected, below LODs, or within lower concentration levels (Table 2; Figure 2).

**Figure 2:**
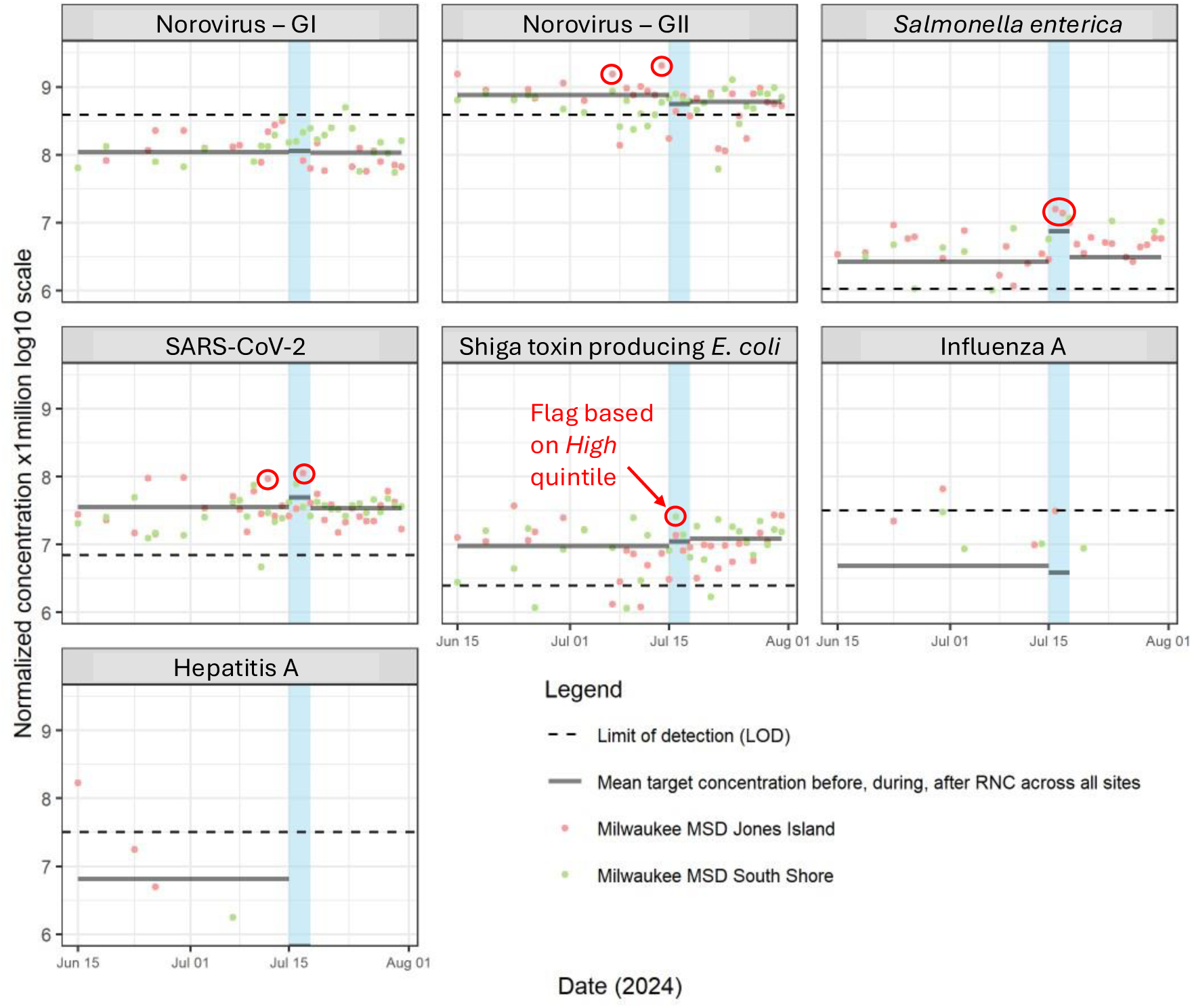
Flow and population normalized wastewater concentration data for various pathogens monitored during the 2024 Republican National Convention – Milwaukee, WI, July 2024. The dates of the 2024 Republican National Convention (RNC) are highlighted in blue. Geometric mean concentration across all sites for before, during, and after the RNC are displayed as horizontal lines across the corresponding time period. The limit of detection (LOD) line represents the mean value of all population-normalized LOD values throughout the entire surveillance period specific to each pathogen. Observations flagged during the event surveillance period are circled in red.

**Table 2:**
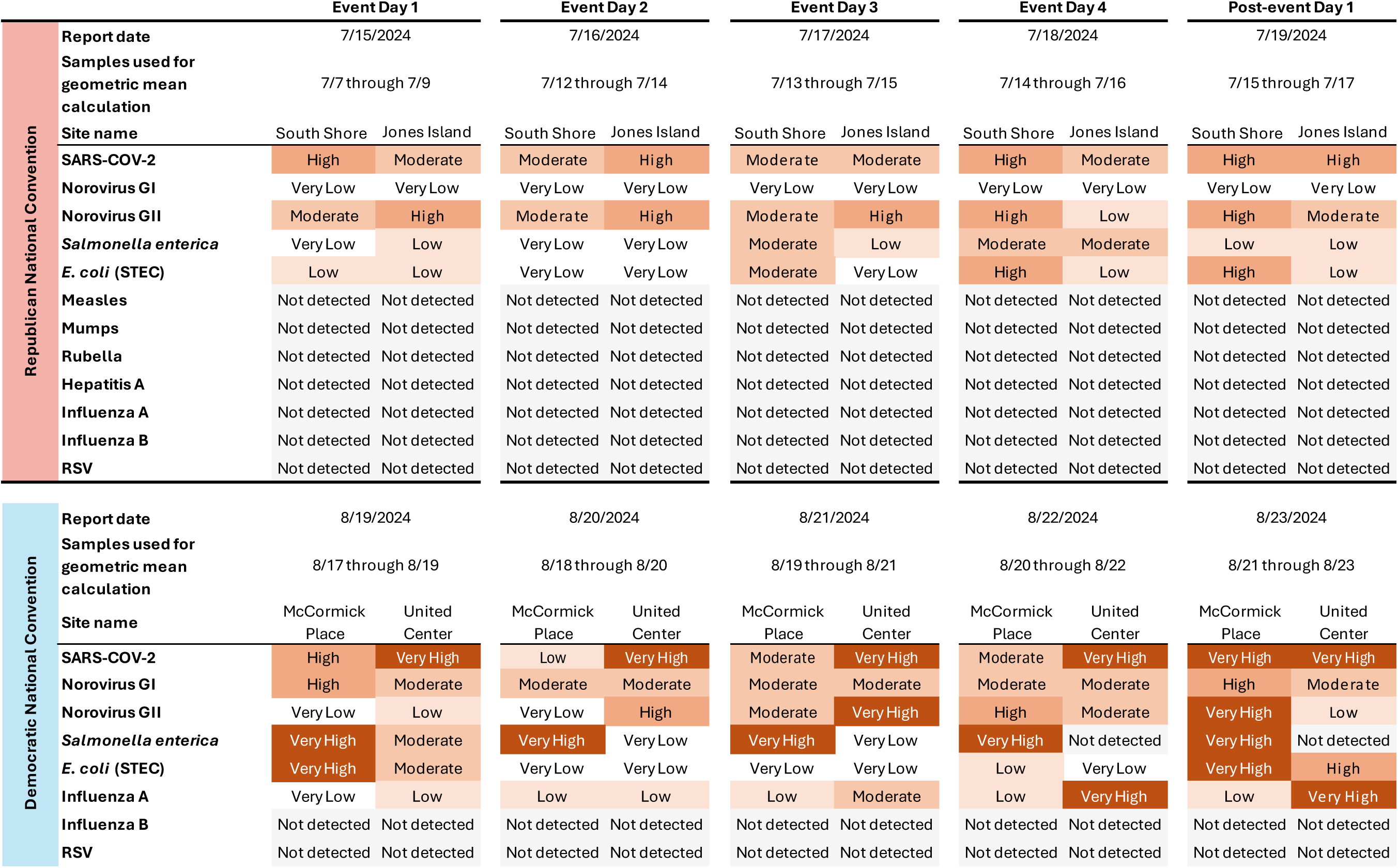
Quintile levels of wastewater concentration data (3-day running average) reported to the surveillance task forces during the 2024 Republican National Convention (Milwaukee, WI, July 2024) and the 2024 Democratic National Convention (Chicago, IL, August 2024).

We observed sporadic detections of HAV and IAV prior to the event, including a significant spike in HAV levels (200,000 gc/L) on June 15 in the Jones Island sewershed. An HAV epidemiologist was immediately consulted but no public health action was ultimately taken because (i) the Milwaukee area had no known contemporaneous cases or outbreaks of HAV and (ii) no subsequent detection above LOD occurred during the monitoring period. The prevailing theory remains WBE discovery of an undiagnosed case or cluster of HAV residing in the sewershed at the time of detection.

### DNC results

During the DNC event, samples results were available within 12 hours of collection. There was more instability in target quintiles recorded during the DNC compared to the RNC, with 17 *Very High* levels recorded among five of the eight targets across the five-day event window (Table 2). These results were anticipated because baseline sampling at McCormick Place and United Center had started only 1.5 months before the DNC response, relative to the >1 year’s worth of baseline data at Milwaukee sites. Therefore, for the DNC, more emphasis was placed on concentration trends over time (Figure 3) and we did not observe concerning increases or spikes in concentration levels for most pathogens. RSV was not detected in wastewater before, during, or after the event; IBV was only detected once, several weeks before the DNC began and IAV was detected sporadically at low levels. SARS-CoV-2 and NoV GI were detected at concentrations similar to those detected pre-event. We noted a rise in NoV GII on the final two days of the DNC which exceeded the maximum observed value during the baseline period. We also observed elevated STEC levels on the final day and the day after the DNC.

**Figure 3:**
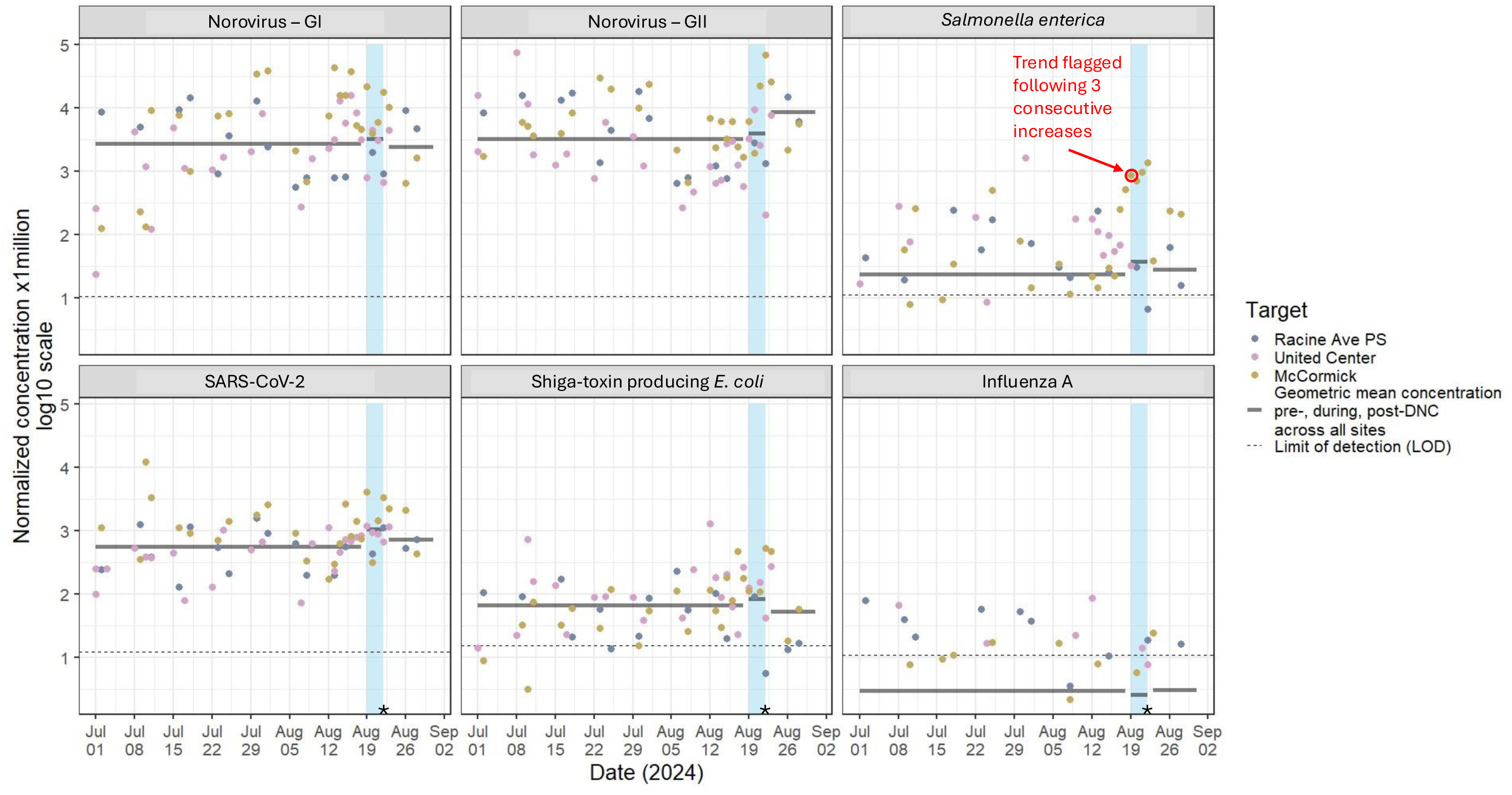
Flow and population normalized wastewater concentration data trends for various pathogens monitored during the 2024 Democratic National Convention – Chicago, IL, August 2024. The dates of the 2024 Democratic National Convention (DNC) are highlighted in blue. Geometric mean concentration for periods before, during, and after the DNC are displayed as horizontal lines across the corresponding time period. The limit of detection (LOD) line represents the mean value of all population-normalized LOD values throughout the entire surveillance period specific to each pathogen. PS = pumping station. *Due to low water levels, a grab sample was collected from the United Center manhole on August 22.

The only pathogen-specific WBE trend that prompted a public health response during the DNC occurred on August 19, 2024—the first day of the convention—when we detected a third consecutive daily increase of normalized *S. enterica* concentration from the manhole nearest the McCormick Place venue (annotated in Figure 3). Epidemiologists then planned for continued monitoring with simultaneous investigation into possible corroborating signals in other syndromic surveillance data sources. Epidemiologists present at the venue first aid stations were advised to monitor any signs of gastrointestinal illnesses. *S. enterica* levels continued to increase throughout the event, eventually peaking on August 22, the last day of the DNC. Despite the substantial and progressive increase in wastewater levels of *S. enterica*, there were no corroborating signals in other data systems; emergency department visits for gastrointestinal symptoms remained below baseline and no medical aid station visits were associated with gastrointestinal illness nor potential *S. enterica* exposure. Furthermore, increased *S. enterica* levels were not seen at the other DNC sampling site (United Center) nor from any other sampling location in Chicago and on August 23, 2024, one day after DNC activity at McCormick Place ended, *S. enterica* levels dropped to pre-event levels. In consideration of WBE trends and the lack of substantiation in other data sources, no public health action was taken.

In an attempt to better understand the WBE signal that was detected during the DNC, all wastewater specimens collected from McCormick Place were selectively cultured. *S. enterica* isolates were cultured from the five McCormick Place daily samples collected from August 18 through August 22 that were associated with increased *S. enterica* levels, no isolates were cultured from other days. These isolates were submitted for whole-genome sequencing by the Regional Innovative Public Health Laboratory (RIPHL), an academic/public health partnership between Rush University Medical Center and CDPH. All five isolates were assembled into high quality genomes and all shared the same multilocus sequence typing (MLST) profile (Supplementary Material available upon request), though the sequence type was undefined on PubMLST (pubmlst.org). All isolates were identified as belonging to the same *S. enterica* serotype Isangi. Upon core genome single nucleotide polymorphism (SNP) analysis, three isolates were identical and the remaining two isolates were 1 and 3 SNPs diverged from the identical isolates. Antimicrobial susceptibility testing indicated that all five isolates were susceptible to ampicillin, ceftriaxone, levofloxacin, and trimethoprim-sulfamethoxazole.

## Discussion

Overall, WBE was readily incorporated into public health pathogen surveillance for both the RNC and DNC in 2024—two large-scale events of significant national importance. A key factor in the successful application of WBE during the response was the pre-existing operational status of the two WBE programs, each with several years of experience supporting wastewater monitoring initiatives. In other words, as noted by the Committee on Community Wastewater-based Infectious Disease Surveillance at the National Academies of Sciences, the maintenance of a “wastewater surveillance system ensures readiness to respond to evolving risks.” ^39^ In the lead-up to the RNC and DNC, extensive collaboration between the Wisconsin and Chicago teams played a critical role in enabling a coordinated launch of large-scale wastewater monitoring for these events. This concerted planning effort, which took place over the course of more than a year, enabled efficient turnaround of data for a panel of eleven target pathogens during the RNC and seven target pathogens during the DNC. WBE application for two of these pathogens (*S. enterica* and STEC) was new to both teams for the purpose of this dedicated response and was included given pathogen relevance to large-scale events.^22,23,36,37^ Jurisdictions considering wastewater surveillance for upcoming large-scale events might consider beginning discussions with their respective WBE programs at least 12 months in advance of the event.

The importance of public health response plans in creating a useful WBE program has been noted previously and this was true for both the RNC and DNC.^40^ In both Milwaukee and Chicago, the advanced planning around the interpretation and utilization of WBE data was instrumental. This included sharing visualization formats (e.g., Table 2, Figures 2 and 3) in advance with convention decision-makers within the state’s Public Health Emergency Preparedness (PHEP) and operations teams in addition to updating them throughout the course of the events. We also established a common public health response plan based on pathogen and site-specific thresholds (summarized in Figure 4) which may be useful to other public health jurisdictions considering WBE for large-scale events. Surveillance of higher consequence pathogens, such as measles and HAV, involved a lower threshold to activation compared with more endemic pathogens; for instance, a single detection above the LOD for measles virus would have been sufficient to warrant subsequent public health investigation and action. Following WBE signal validation and contextualization, we specified the need for follow-up including possible public messaging, alerts to healthcare providers, or other guidance for event organizers based on the pathogen and threat level (Figure 4).

**Figure 4:**
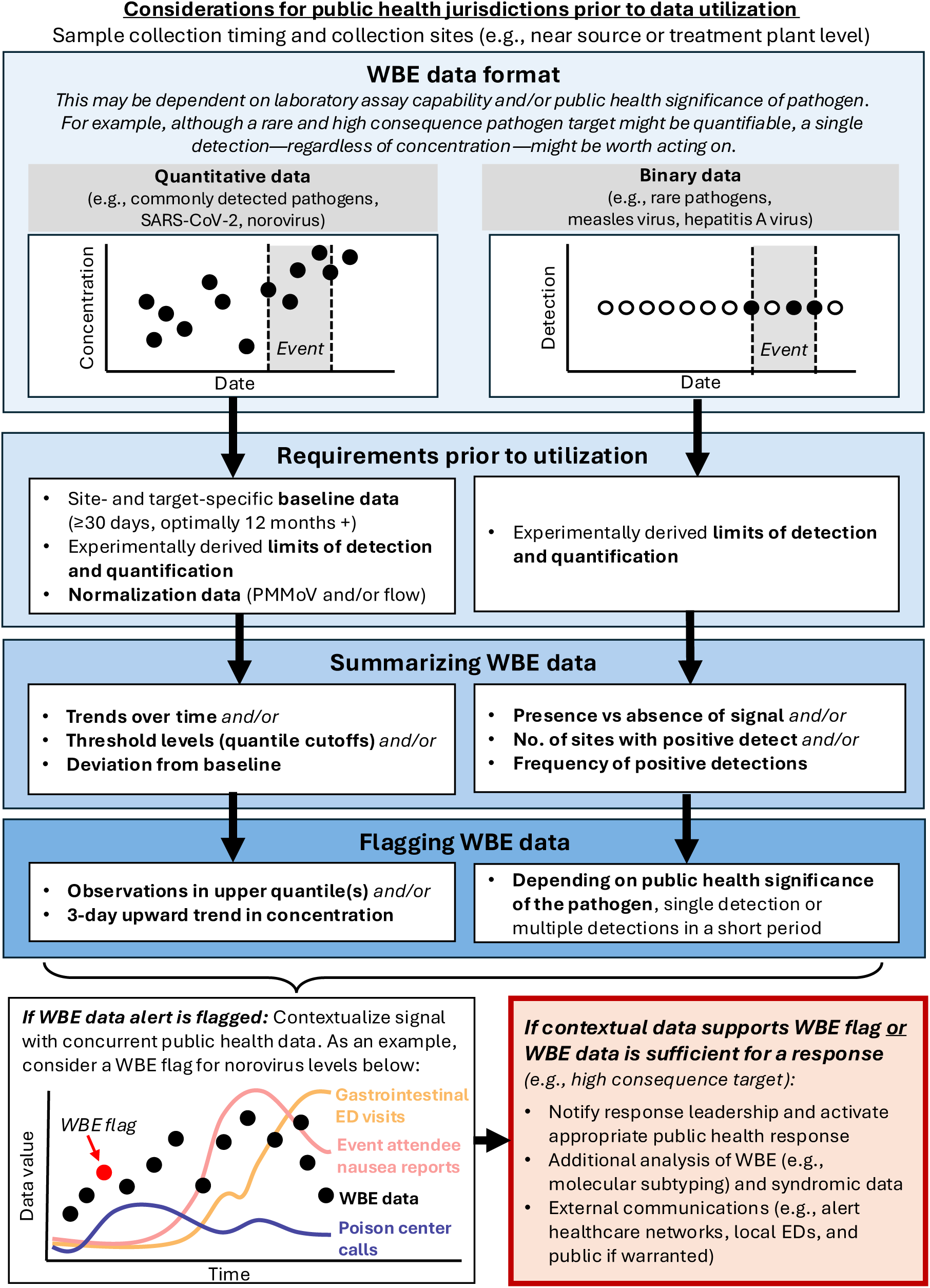
Recommended response framework for wastewater-based epidemiology at large-scale events.

A large component of interpreting WBE data was centered on contextualization, especially for pathogens regularly found in wastewater. The baseline data collected before the RNC and DNC events were essential in defining thresholds for action. Past WBE work reinforces this idea: a 2003 norovirus outbreak on a university campus was recognized in WBE data based on its sharp contrast to an extended six-week baseline period, after which point public health intervened with an enhanced cleaning protocol and a public information campaign.^41^ Without sufficient baseline data to compare against, pathogen fluctuations during a time period of interest can be challenging to interpret. Established sewersheds with substantial sample history can provide for a more robust alerting framework and one that is less susceptible to the frequent large deviations in WBE data (which are even more common in smaller sewersheds). Indeed, the value of extended baseline sampling was evident in our findings. Quintile category fluctuations (essentially the magnitude of deviation within the baseline distribution) were notably less pronounced in Wisconsin, where sampling had occurred at larger WRFs for over a year, compared to Chicago where sampling from smaller sewersheds began only 1.5 months prior to the event. An ideal baseline would reflect data collected during the same calendar period of surveillance as the event is occurring (from the year or years prior) in order to compare against any seasonal trends. At the same time, while collection of longer baseline periods might be ideal, this recommendation should be considered alongside cost-efficiency and the resources need for extended WBE sampling and processing.^39^

Just as baseline wastewater data assisted with internal contextualization, we found it equally important to interpret WBE data side-by-side with other public health data sources (e.g., syndromic surveillance from clinical settings, laboratory clinical test results, ambulance run data, on-site first aid station visits). Utilizing WBE at both the RNC and DNC benefited from the existence of other jurisdictional public health surveillance systems because, as Nkambule *et al* noted, “wastewater data should not be interpreted in isolation, but rather used in conjunction with other health data”.^33^ The acute increases in *S. enterica* concentration observed during both the DNC and RNC are an excellent testament as to why. Absent corroboration from any other surveillance system, the *S. enterica* levels detected in Chicago far exceeded baseline data levels (and less significantly so in Wisconsin) and might have prompted public health action such as increased food safety inspections and/or public announcements at the associated venue. These would all have been decisions with resource, public notification, and media coverage implications. Instead, there was no substantiating data in any other surveillance source (e.g., no nausea or diarrhea complaints reported to medical tents, no notable change in emergency department visits associated with gastrointestinal illness). Ultimately, no extensive public health response was taken.

The definitive cause of the *S. enterica* spike at the DNC and RNC remains unknown but was in hindsight, perhaps unsurprising. First, *S. enterica* is common to mass gatherings.^22,23^ Second, the bacteria is frequently detected in wastewater absent corroborating clinical data, such as in Japan from 2016–2019^42^ and in Hawaii in 2011,^9^ though these detections occurred over the course of months and years rather than a 5-day detection period such as ours. At both conventions, we may have detected an *S. enterica* signal from at least one subclinical case in a person within the sewershed, convention-related or not. We also considered the possibility of persistent raw food contamination associated with food preparation practices at the venues. As more pathogen targets are added to WBE programs and our understanding of WBE utility for different pathogens expands, we may ultimately find that elevated WBE signals for pathogens like *S. enterica* are sufficient to initiate public health action even in the absence of clinical corroboration. It is also true that had we detected a more consequential pathogen, like measles virus rather than *S. enterica*, the burden of evidence needed for public health action would have been considerably lower.

For public health jurisdictions that may be considering WBE for large-scale events, we offer several considerations. One important consideration is that during the event response meetings which were established for both conventions, many different programs, experts, and skillsets were present in the same room. To avoid misinterpretation of WBE data by stakeholders and decision makers, WBE programs should make a concerted effort to establish the purpose and limitations of each surveillance system utilized, including WBE (given what we know about WBE to date). We framed WBE as a complementary tool within the public health surveillance toolbelt, which is known to correlate with traditional clinical surveillance data for many pathogens, including SARS-CoV-2,^16,30,43^ IAV,^15,16,18,44^ RSV,^15^ *S. enterica,*^10^ and NoV.^17^ We highlighted its unique characteristics in the context of surveillance at large-scale events—namely, its ability to capture data from broad populations and its ability to detect disease signals prior to traditional clinical surveillance data. At the same time, we also described its limitations—specifically, pathogen-specific shedding dynamics and viral durability in wastewater which may influence the timing and magnitude of WBE data patterns. This is particularly pertinent to surveillance at large-scale events, which may only last several days. WBE can only detect pathogens shed from infected individuals and, for an infection with a long incubation period, these signals might only appear in wastewater after the conclusion of a short event. For this reason, the WWMP and CWBE programs extended surveillance for multiple weeks after the conclusion of the events.

A second important takeaway is that epidemiologists should consider how timing of sample collection may influence results, especially when using Moore swabs (as was done in Chicago). Although gauze-like samplers have been shown to provide WBE data comparable to composite sampling,^45,46^ the sampler’s saturation may limit uptake of pathogens after some extent of time; one field study in Canada found linear accumulation of SARS-CoV-2 material for up to 48 hours in gauze membranes,^47^ while other work suggests that suspended solids may saturate Moore swabs within 8 hours.^48,49^ At the DNC, Moore swabs were placed in manholes at 9:00am each day and left for 24 hours. As a result, swabs at the McCormick Place manhole, where attendees conducted Convention business from 9:00 am to 5:00 pm, may have more appropriately captured wastewater contributions than its counterpart at the United Center site, where events only began at 8:00pm each night. If a similar event were held in the future, CDPH would reconsider when samplers were deployed at specific sites based on when each venue was fully occupied. Composite sampling (as performed in Milwaukee, Wisconsin) could have effectively addressed this issue; however, city restrictions prevented autosamplers from being deployed inside manholes.

An additional factor to consider is the location of sample collection points. For one, WBE programs should consider the physical distance from the collection site to the processing laboratory; in our experience, this was our primary limiting factor in turnaround time. Whereas Chicago samples were collected within several kilometers of the UIC laboratory, Milwaukee samples were transported >110 km to the WSLH to assess for 8 of the 11 pathogens monitored. Correspondingly, while samples for the DNC could be resulted in <12 hours, RNC sample results were available to WWMP in closer to 36 hours from collection. Jurisdictions in need of same-day WBE results (particularly relevant to large-scale events of short duration) might consider enlisting the help of a local laboratory to test for most, if not all, of the target pathogens. Second, the specific choice of sampling sites should consider the goals of the surveillance program and event details. For more targeted attribution and public health responsiveness during large-scale events, jurisdictions might be interested in conducting sampling closer to the event venue (as was performed in Chicago).^50,51^ The closer a WBE sample is collected to the source of interest, the more actionable it may become.^35,41,52,53^ Though concentration values can be noisier, sampling at the “sub-sewershed” level also has the distinct advantage of increased sensitivity, where pathogen targets are more readily identified during periods of little-to-no transmission.^52^ This is possibly evidenced by repeated detections of influenza A virus in Chicago but not in Milwaukee, despite both conventions occurring during traditionally low-transmission months prior to the early-October start of respiratory season. It is important to note that establishing these new sites involved several months of discussions, documentation, and approvals from both local and federal authorities. Conversely, community-level sampling at water reclamation facilities, as conducted in Milwaukee, offered potential advantages including: (i) more stable and representative baseline data, (ii) broader catchment coverage for identifying event-associated threats, and (iii) the ability to detect spillover into surrounding communities—an aspect that may be missed when monitoring is limited to areas in or near event venues. Ideally, if resources permit, samples should be collected both from community-scale treatment plants—to capture trends within the broader population—and from locations proximal to the event venue. This dual approach enables public health officials to determine whether a disease signal is widespread or localized to the event-specific area.

Ultimately, we successfully integrated WBE into public health surveillance protocols for mass gatherings, providing each jurisdiction with rapid, pathogen-specific data, cost-effective population-level sampling, and complementary insights to augment existing syndromic surveillance systems. Much of our success in incorporating WBE into the established public health surveillance protocol was due to advanced planning by both laboratorians, epidemiologists, and other public health practitioners across two US cities; this inter-jurisdictional collaboration was essential in sharing best practices, aligning priorities, and corroborating results. Although we were limited in our ability to evaluate the full potential of WBE during large-scale events, given the absence of major public health threats during either convention, we hope that our experience might provide some guidance to other public health jurisdictions considering WBE for large-scale events. This may be pertinent given several high-profile, upcoming events in the United States, including the 2026 FIFA World Cup and the 2028 Summer Olympics and Paralympic Games in Los Angeles, California. With sufficient planning and by adopting, modifying, standardizing, and improving upon methods used at the RNC and DNC, local public health officials can rely on WBE data to provide a more comprehensive understanding of health threats during consequential large-scale events.

## Data Availability

All data produced in the present study are available upon reasonable request to the authors.

## Acknowledgements

*Chicago Department of Public Health:* Dorothy Foulkes, Kendall Anderson, Chris Shields, Janna Kerins and the Communicable Diseases Program; *Current*: Melissa Pierce, Aamer Ahmed; *University of Illinois Chicago:* Mayumi Pascual and Michael Secreto; *Wisconsin Department of Health Services Communicable Disease Epidemiology Staff; Milwaukee Metropolitan Sewerage District Staff; Wisconsin Wastewater Surveillance Program staff at the Wisconsin State Laboratory of Hygiene*: Erica Camarato, Evelyn Doolittle, Devin Everett, Jocelyn Hemming, Suzanne Joneson, Griffin Knuth, Oliver Long, Ashlie McCunn, Shreya Shrestha, Laura Simonson, Regan Wied; *Regional Innovative Public Health Laboratory:* Ellen Gough; *Wisconsin Wastewater Surveillance Program staff at the University of Wisconsin-Milwaukee*: Kieyarrah Dennis, Benjamin Gallion, Angela Schmoldt, Melinda Bootsma; *City of Milwaukee Health Department:* Ryan Honeck, Sivani Manchu

